# Development and Reliability of an Objective Structured Clinical Examination for Intake Interviews in Mental Health Occupational Therapy Education

**DOI:** 10.1101/2025.11.22.25340285

**Authors:** Yasuhisa Nakamura, Shohei Mori, Shuji Kijima, Masahiro Tanaka, Osamu Taguchi

**Affiliations:** Department of Rehabilitation, Division of Occupational Therapy, Faculty of Health Sciences, Nihon Fukushi University, Handa City, Aichi Prefecture, Japan; Faculty of Rehabilitation, School of Health Sciences, Fujita Health University, Toyoake, City, Aichi Prefecture, Japan

## Abstract

**Introduction:** Evaluating clinical competencies in mental health occupational therapy is hampered by the lack of standardized assessment tools. Intake interviews are particularly difficult to assess because of their interpersonal and observational nature. This study developed an Objective Structured Clinical Examination (OSCE) for intake interviews in mental health occupational therapy education and examined its reliability.

**Methods:** In this cross-sectional study, 60 third-year occupational therapy students from two Japanese universities each completed one OSCE station using one of three standardized patient scenarios based on common psychiatric presentations. Performance was rated on a 16-item, behaviorally anchored scale across three domains (attitudes, skills, evaluation) by two occupational therapists. Internal consistency was assessed using Cronbach’s alpha, and inter-rater reliability using intraclass correlation coefficients; agreement on total scores was examined with Bland–Altman analysis.

**Results:** Internal consistency was high for all domains (Cronbach’s α = 0.850–0.887) and for the total scores (α = 0.816 and 0.817). Inter-rater reliability ranged from moderate to excellent across domains (ICC = 0.670–0.900) and was substantial for the total score (ICC = 0.854, p < 0.001). Bland–Altman analysis showed a small mean difference between raters and narrow 95% limits of agreement.

**Discussion:** This OSCE showed high internal consistency and acceptable inter-rater agreement in assessing complex competencies such as empathy and professionalism. By translating abstract interpersonal skills into observable behaviors, it may provide a practical framework for structured formative feedback. Further studies with larger and more diverse samples are needed to confirm its validity and applicability.

## Introduction

Objective Structured Clinical Examinations (OSCEs) are widely used to assess clinical competencies in healthcare education, particularly in medical and nursing programs [1,2]. These assessments typically employ standardized patients (SPs) to evaluate practical skills such as communication, interviewing, and clinical reasoning [3,4]. Although OSCEs have also been introduced in occupational therapy education, their use in mental health contexts remains limited, especially with respect to structured and psychometrically evaluated tools [5,6].

In mental health occupational therapy, essential competencies such as empathetic communication, appropriate nonverbal behavior, and professional demeanor are difficult to assess using traditional written or knowledge-based examinations [7,8]. Clinical reasoning in mental health is complex and context dependent, further complicating standardized assessment. As a result, educational assessments in this field often overemphasize theoretical knowledge while underrepresenting practical and interpersonal skills that are central to psychiatric care [9].

Some studies have reported the use of OSCEs in occupational therapy; however, few have empirically evaluated tools specifically designed for mental health settings. In particular, assessments of psychiatric intake interviews frequently rely on global or subjective impressions, which reduces consistency and transparency [10,11]. Thus, although there is broad recognition of the need to evaluate relational and observational skills, standardized tools with documented psychometric properties remain scarce [6].

To address this gap, the present study adopted an educational assessment approach that emphasizes behavioral anchoring, namely the use of observable indicators to evaluate complex clinical skills. Prior work suggests that operationalized behavioral criteria can enhance inter-rater reliability and support more concrete formative feedback [12,13]. Such criteria may also improve measurement precision and foster a shared evaluative language between educators and students [14].

This study therefore aimed to develop an OSCE tailored to intake interviews in mental health occupational therapy and to examine its reliability, focusing on internal consistency and inter-rater agreement. Using three SP scenarios representing common psychiatric presentations, student performance was rated with a 16-item, behaviorally anchored scale. Statistical analyses were conducted to evaluate internal consistency and inter-rater reliability. This initial psychometric evaluation is intended to provide a foundation for a structured OSCE framework in psychiatric occupational therapy education, while recognizing that further research is needed to establish its validity and educational impact.

## Materials and Methods

### Study design and setting

This cross-sectional observational study was conducted between September 2024 and July 2025 at two Japanese universities that offer accredited occupational therapy programs. The OSCE was implemented as part of a mandatory educational module for mental health occupational therapy. All assessments were conducted in clinical skills training rooms designed for simulation-based education.

The OSCE followed a single-station format. Each station consisted of one intake interview with an SP, followed by a 5-min period in which the raters completed the scoring sheet.

### Participants

Sixty third-year undergraduate occupational therapy students participated in the OSCE, with 30 students from each university. All students had completed the required coursework in mental health and were preparing for their clinical placements. Participation in the OSCE was compulsory as part of the course requirements. Students who were absent or failed to complete the full assessment were excluded from the analysis. Written informed consent was obtained from all participants. The study was approved by the Ethics Committee of Nihon Fukushi University (Approval No. 23-042-02).

### OSCE development

The OSCE was developed to provide a structured and behaviorally anchored method for assessing practical competencies that are often underrepresented in written or knowledge-based assessments, particularly in mental health occupational therapy. Item construction was informed by a literature review on psychiatric interviewing, occupational therapy education, and prior OSCE frameworks [5,6,9].

An initial pool of items relevant to intake interviews was drafted and then refined through multiple expert panel discussions involving occupational therapy educators specializing in mental health. The panel included educators and clinicians with experience in psychiatric occupational therapy; their professional backgrounds and roles are summarized in Supplementary Table 2. Through iterative discussion, the panel qualitatively reviewed item wording, domain allocation, and behavioral anchors until consensus was reached. No formal content validity index was calculated; instead, the initial version of the scale was established through this qualitative consensus process.

The final version of the rating scale comprised 16 items across three domains: attitude (four items), skills (nine items), and evaluation (three items). Item categorization was based on theoretical frameworks of interpersonal skills and observable clinical behaviors. For example, “Seating and distance” was categorized under “Skills” rather than “Attitude” because it involves explicit, observable nonverbal behavior. According to Sherer and Rogers [15], nonverbal behaviors such as physical distance and seating arrangement represent practical skills in therapeutic communication rather than merely reflecting an inner disposition.

Each item was rated on a three-point scale (0 = not demonstrated, 1 = partially demonstrated, 2 = fully demonstrated), yielding a total score ranging from 0 to 32. All items were behaviorally anchored to increase objectivity and minimize subjectivity in rater judgment [12,13]. The complete rating criteria are presented in Supplementary Table 1. The rating scale was piloted in 2023 with a separate cohort of 20 second-year occupational therapy students from one of the participating universities. Participants in the pilot provided feedback on the clarity, feasibility, and usability of the scale. Based on this feedback, item wording—particularly within the skills domain—was simplified, and several behavioral anchors were refined to improve precision and ease of use.

### Procedure

Each student completed one OSCE session using a single SP scenario. Students were informed that three different scenarios could be presented, but they did not know in advance which scenario they would encounter. Students were randomly assigned to one of the three cases using a random number table. To avoid imbalance between universities, the number of students assigned to each case was capped at 10 per university, resulting in approximately equal distributions of scenarios across the two sites.

All scenarios reflected common psychiatric presentations associated with moderate functional impairment (Global Assessment of Functioning score of approximately 40) and were designed to simulate realistic inpatient settings:

- Scenario 1: auditory hallucinations, insomnia, and irritability
- Scenario 2: persecutory delusions and somatic preoccupations
- Scenario 3: negative symptoms such as avolition and emotional fatigue

The SPs were undergraduate students who did not belong to the target cohort. They received scenario-specific training under faculty supervision. Training included viewing video examples of intake interviews for each scenario, rehearsing the roles, and receiving feedback to ensure consistency in symptom presentation and interaction style across sessions. This approach is consistent with recent studies supporting the reliability and authenticity of student SPs [16].

Each simulated interview was observed and independently scored by two licensed occupational therapists, each with more than 10 years of clinical experience in mental health. The two raters evaluated all 60 students. They were aware of the students’ university of origin and assigned scenario; thus, blinding was not implemented. Interviews were video recorded, and the recordings were available for review when raters needed to confirm their real-time judgments. The raters used the 16-item OSCE scale and scored each item in real time based on the predefined behavioral anchors.

Before data collection, the raters underwent standardized training that included familiarization with the scoring tool, joint review of pilot OSCE videos, and consensus-building exercises to align their interpretations of each behavioral anchor [17]. Calibration meetings were also held during the study period to minimize rater drift and maintain consistency in scoring.

## Statistical analysis

Descriptive statistics (means, standard deviations, medians, and ranges) were calculated for the attitude, skills, and evaluation domains and for total scores from both raters.

Internal consistency was examined using Cronbach’s alpha for each domain and for the total score for each rater, with α ≥ 0.80 considered acceptable.

Inter-rater reliability was evaluated using intraclass correlation coefficients (ICCs) based on a two-way random-effects model with absolute agreement, single measures [ICC(2,1)]. ICCs were calculated for each domain and the total score, and 95% confidence intervals were reported. ICC values were interpreted according to conventional benchmarks: moderate (0.60–0.74), substantial (0.75–0.89), and excellent (≥ 0.90).

To further assess agreement between raters for the total score, Bland–Altman analysis was conducted. The mean difference and standard deviation (SD) of the differences were calculated, and the 95% limits of agreement (LOA) were derived as mean difference ± 1.96 × SD. Fixed bias was tested using a one-sample t-test of the mean difference against zero (df = 59), and proportional bias was examined using simple linear regression with the mean total score as the independent variable and the score difference between raters as the dependent variable.

All analyses were conducted using IBM SPSS Statistics for Windows, Version 21.0, with the significance level set at p < 0.05.

## Results

### Descriptive statistics and inter-rater reliability

The internal consistency of the OSCE rating scale was high across all domains and raters. Cronbach’s alpha ranged from 0.850 to 0.887 for each domain and rater, and the total scores showed values of 0.816 and 0.817, respectively. These findings indicate strong internal coherence of the scale across multiple components. The full reliability coefficients are presented in Table 1.

**Table 1.**
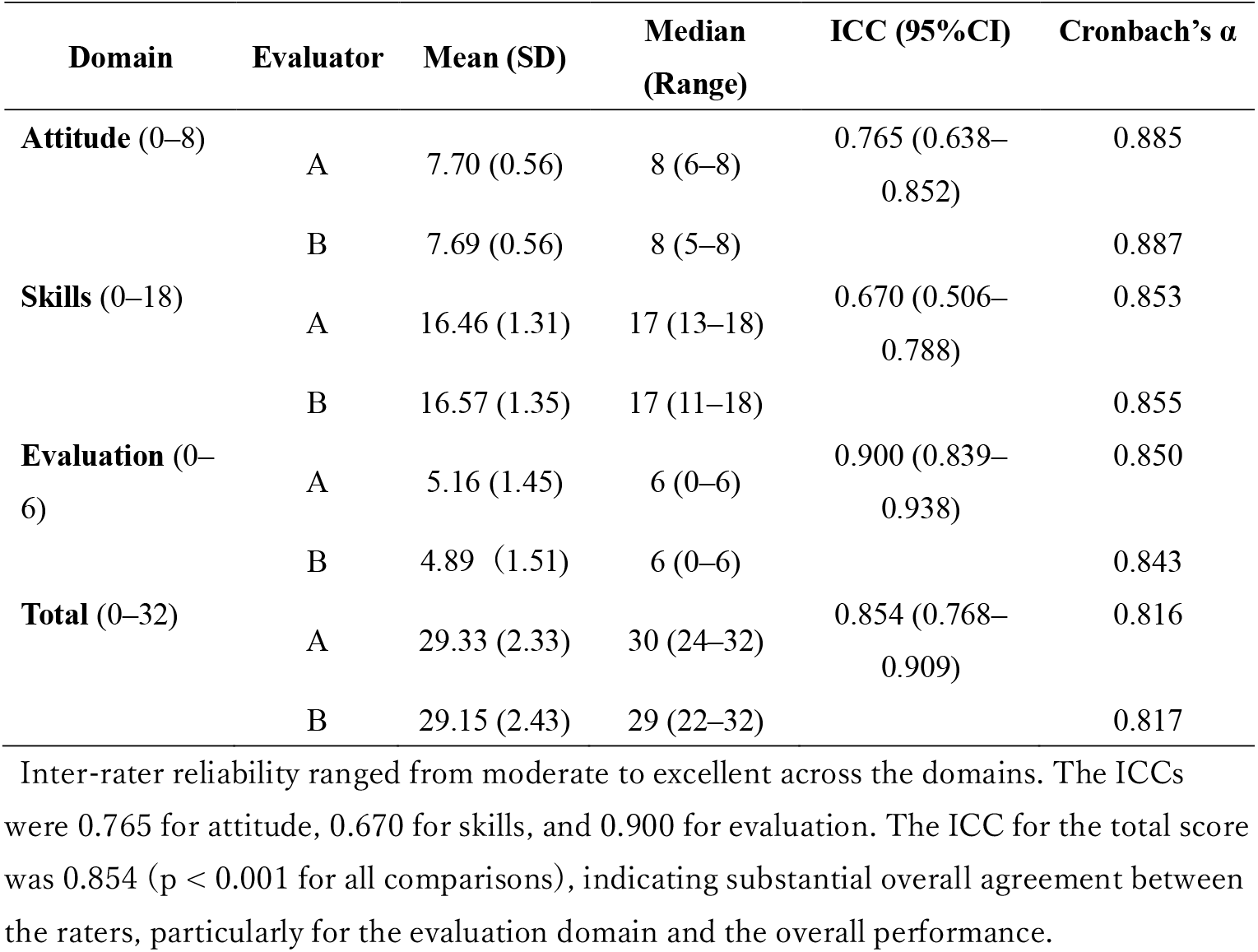
Descriptive statistics, inter-rater reliability (ICC), and internal consistency (Cronbach’s α) of OSCE domains.

Scores were generally high across all three domains. For the attitude domain, both raters assigned similar scores (mean = 7.70 and 7.69; median = 8). The skills domain showed slightly more variability (mean = 16.46 and 16.57), while the evaluation domain had the widest range (mean = 5.16 and 4.89). The total scores were consistently high across raters (mean = 29.33 and 29.15). Table 1 summarizes the descriptive statistics for each domain.

### Inter-rater agreement visualization

A Bland–Altman plot was used to visualize agreement between the two raters’ total OSCE scores (Figure 1). The mean difference between raters was small (0.18, SD = 1.29), with 95% limits of agreement ranging from −2.35 to 2.71. Most data points fell within these limits, indicating acceptable agreement between the two raters for the total OSCE scores.

**Figure 1.**
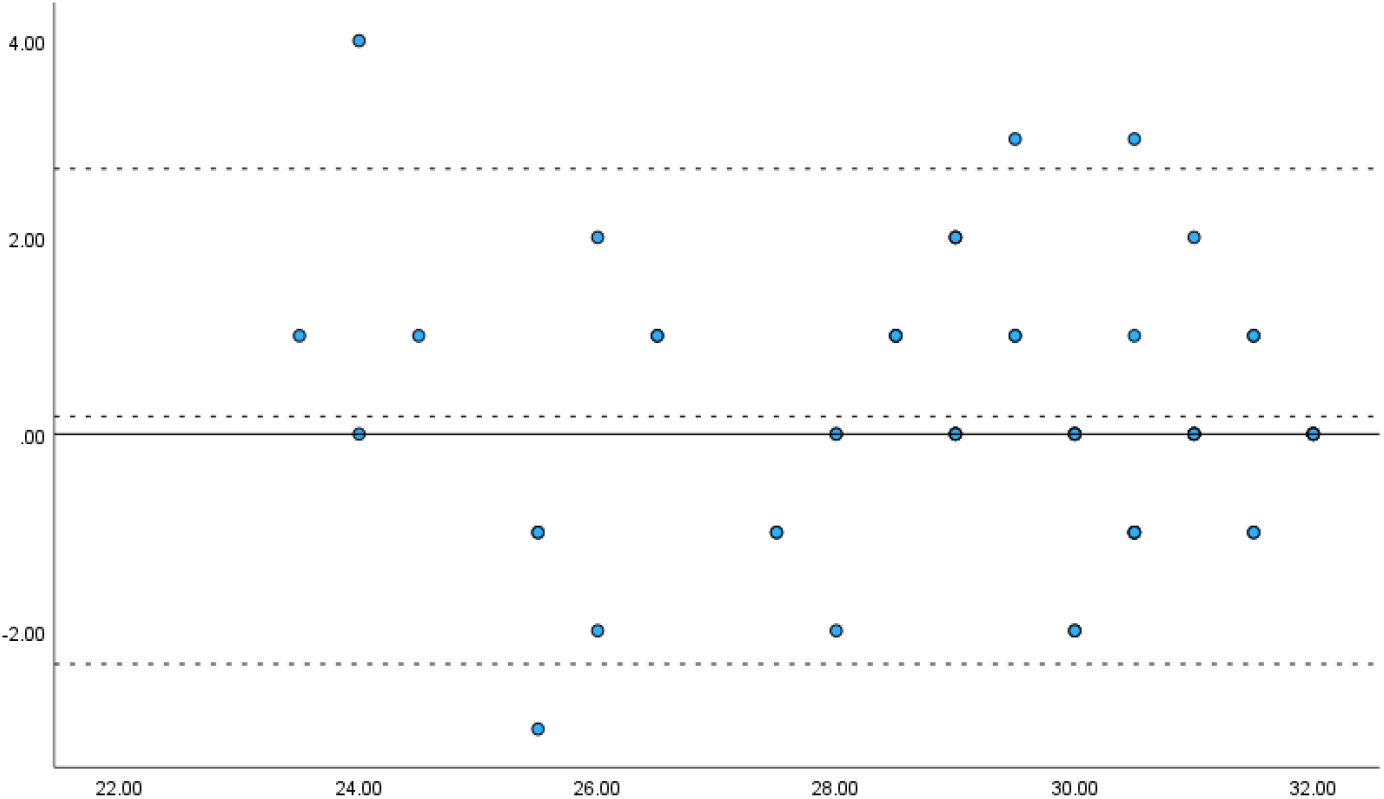
Bland–Altman plot of inter-rater agreement for total OSCE scores (n = 60). Note. The solid line indicates the mean difference between raters (0.18), and the dashed lines indicate the 95% limits of agreement (−2.35 to 2.71). X-axis: mean of the two raters’ total scores; Y-axis: score difference (Rater A − Rater B).

### Fixed and proportional bias

A one-sample t-test indicated that the mean score difference between raters was not significantly different from zero (t(59) = 1.096, p = 0.277), indicating no fixed bias. The effect size was small (Cohen’s d = 0.14), and linear regression analysis revealed no significant proportional bias (*β* = −0.045, p = 0.540). These findings support the reliability of the scoring process and the absence of systematic rater bias.

## Discussion

This study developed and conducted an initial psychometric evaluation of an OSCE specifically tailored to intake interviews in mental health occupational therapy education. The findings indicated high internal consistency, substantial to excellent inter-rater reliability, and good scoring reproducibility in this sample, suggesting that this OSCE may provide a useful structured option for assessing complex clinical competencies within this educational context. Because this initial study focused on reliability indices, other aspects of validity—such as structural and criterion-related validity—were not directly examined and remain important targets for future research.

First, internal consistency was high across all domains. Cronbach’s alpha values ranged from 0.850 to 0.887, and the total score showed similarly strong internal consistency (Cronbach’s *α* = 0.816–0.817). These results indicate that the OSCE items are coherently organized and capture related facets of clinical competency within each domain.

In addition, inter-rater reliability was substantial to excellent, providing evidence of generally consistent scoring between raters. Specifically, ICC values were 0.765 for the attitude domain, 0.670 for skills, and 0.900 for the evaluation domain. The total score ICC (0.854, p < 0.001) also supported good overall reliability. The particularly high agreement in the evaluation domain likely reflects its emphasis on well-defined, observable behaviors such as summarizing patient content and identifying speech patterns. By contrast, the comparatively lower ICC in the skills domain is consistent with prior research, which has highlighted the inherently interpretive nature of rating interpersonal aspects such as empathy, neutrality, and professionalism [17,18]. Further refinement of behavioral anchors, together with structured rater training using standardized video exemplars, may help enhance reliability in these more subjective areas.

Moreover, Bland–Altman analysis supported the consistency of the rating process. The mean difference between raters was small (0.18, SD = 1.29), and there was no evidence of meaningful fixed bias (p = 0.277, Cohen’s d = 0.14) or proportional bias (β = −0.045, p = 0.540). Taken together with the ICC results, these findings suggest that the scoring system can generate reasonably stable ratings across raters.

A key feature of this OSCE is its use of behavioral anchoring. By translating abstract clinical competencies—such as empathy, nonverbal communication, and professional demeanor—into explicitly observable and measurable actions, the tool has the potential to reduce subjectivity in scoring. This is particularly relevant in psychiatric settings, where global and impressionistic judgments have traditionally complicated evaluation [19]. Previous research has shown that clearly defined behavioral indicators can improve inter-rater agreement in OSCEs across health professions, supporting the usefulness of this approach [12,13].

Beyond its measurement properties, the OSCE framework may offer educational benefits. It provides shared, concrete language for structured feedback, which can facilitate reflective and constructive dialogue between students and instructors. The use of realistic SP scenarios can also promote experiential learning and support the development of professional identity within a psychologically safe learning environment. Prior studies have emphasized that structured, behaviorally anchored feedback is important for fostering practical skills and professional behaviors [10,14]. However, these potential educational benefits were not directly measured in the present study and should be examined in future research.

Future research with larger and more diverse samples is needed to examine the structural validity of the scale through exploratory and confirmatory factor analyses. In addition, future studies should examine criterion-related validity by comparing OSCE scores with external measures of communication competence that are already used in occupational therapy education [10].

Continued refinement of behavioral indicators and the development of systematic rater training protocols will be important for further strengthening the reliability and generalizability of this OSCE for broader educational and clinical use.

### Limitations

Despite these strengths, this study had several important limitations. First, the sample was limited to third-year students from two institutions, which may restrict the generalizability of the findings. Future research should involve multiple educational institutions, different academic levels, and diverse geographic regions to strengthen external validity.

Second, although the three OSCE scenarios were designed to reflect common psychiatric presentations, they do not cover the full diversity of mental health conditions. Developing and testing a broader set of scenarios representing a wider range of diagnoses, symptom profiles, and care settings will be important to enhance the breadth of the OSCE.

Third, students’ performance scores were generally high and clustered toward the upper end of the scale, suggesting potential ceiling effects and a restricted score range. This pattern may have inflated some of the reliability estimates and limited the ability of the OSCE to discriminate between different performance levels.

Fourth, the standardized patients were undergraduate students rather than trained actors or real patients. Although student SPs can be a practical and feasible option, their use may have limited the realism of patient interactions and thus may affect the extent to which the findings can be generalized to real clinical encounters. In addition, the raters were aware of the students’ institution and assigned scenario, and subtle expectation effects cannot be completely ruled out.

Finally, this initial evaluation focused primarily on reliability indices. Other aspects of validity, including structural and criterion-related validity as well as direct educational outcomes, were not assessed and should be addressed in future studies.

These limitations highlight the need for ongoing refinement of the OSCE and multisite validation to ensure that the tool remains psychometrically sound and educationally relevant across diverse learner populations, scenarios, and clinical settings.

## Conclusions

This study developed and evaluated an OSCE rating scale specifically designed for intake interviews in mental health occupational therapy. The scale demonstrated strong internal consistency and high inter-rater reliability, indicating its potential as a reliable tool for assessing core clinical competencies in psychiatric settings.

By translating abstract skills, such as empathy, professional demeanor, and clinical observation, into observable behaviors, the OSCE enables structured, objective, and reproducible assessments. These features support immediate educational utility and facilitate standardized training across institutions. However, the relatively lower inter-rater agreement observed in the skills domain highlights the importance of ongoing refinement of behavioral indicators and targeted rater training.

Overall, the OSCE provides a practical foundation for improving clinical education in mental health occupational therapy. Future studies should further explore the broader applicability of this OSCE and its impact on student learning outcomes.

## Supporting information

Structure and scoring criteria of the OSCE for intake interviews in mental health occupational therapy

Demographic and professional characteristics of the expert panel involved in the development of the OSCE rating scale

## Acknowledgments

This study was conducted with the cooperation of the students, faculty, and staff of Nihon Fukushi University and Fujita Health University. The authors express their sincere appreciation for this support. Additionally, the authors are deeply grateful to Professor Takehiko Yamanaka of Nihon Fukushi University for providing the research topic and valuable input.

## Artificial Intelligence use

The authors used ChatGPT (OpenAI) solely for language suggestions and grammar checks during manuscript preparation. All scientific content, analysis, and interpretation were produced by the authors.

## Ethics approval

The study protocol was approved by the Clinical Research Ethics Committee of Nihon Fukushi University, Japan (Approval No. 23-042-02). This study was conducted in accordance with the ethical standards outlined in the Declaration of Helsinki and the Japanese National Guidelines for Clinical Research.

## Data availability

The datasets generated and analysed during the current study are not publicly available because they contain identifiable educational performance data of students and are subject to institutional privacy regulations, but are available from the corresponding author on reasonable request.

