## Supplementary material for "Development and Reliability of an Objective Structured Clinical Examination for Intake Interviews in Mental Health Occupational Therapy Education": Structure and scoring criteria of the OSCE for intake interviews in mental health occupational therapy

**Supplementary Table 1. Structure and scoring criteria of the OSCE for intake interviews in mental health occupational therapy**

| No. | Category | Item | Description | Scoring Criteria (0–2 pts) |
|---|---|---|---|---|
| 1 | Attitude | Greeting and self-introduction | Appropriate attire, clear greeting, and self-introduction | 2 = All 3 fulfilled<br>1 = 1 missed<br>0 = ≥2 missed |
| 2 | Attitude | Patient identification | Verification using at least two identifiers (e.g., name, DOB, and ID) | 2 = ≥2 identifiers<br>1 = 1 identifier<br>0 = None |
| 3 | Attitude | Explanation of the purpose and duration | Explains the purpose, content, and approximate duration of the interview | 2 = All 3 explained<br>1 = 1 missed<br>0 = ≥2 missed |
| 4 | Attitude | Obtaining consent | Informs the patient about the interview and note-taking, and obtains consent | 2 = Both items explained<br>1 = 1 only<br>0 = None |
| 5 | Skills | Seating and distance | Guides patients to their seats with appropriate distance and 90° angle; the patient sits first | 2 = All 3 fulfilled<br>1 = 1 missed<br>0 = ≥2 missed |
| 6 | Skills | Verbal clarity | Maintains appropriate language, pace, and volume throughout | 2 = All 3 maintained<br>1 = 1 missed<br>0 = ≥2 missed |
| 7 | Skills | Non-verbal behavior | Maintains proper facial expressions, gestures, and posture | 2 = All 3 maintained<br>1 = 1 missed<br>0 = ≥2 missed |
| 8 | Skills | Active listening | Uses appropriate listening techniques (e.g., nodding and echoing) | 2 = Appropriate use throughout<br>1 = 1–2 inappropriate uses<br>0 = ≥3 inappropriate uses |
| 9 | Skills | Open-ended questioning | Uses open-ended questions to identify main complaints and needs | 2 = Both identified<br>1 = 1 only<br>0 = None |

| | | | | |
|---|---|---|---|---|
| 10 | Skills | Empathy and neutrality | Shows empathy without denying or affirming the patient's experiences | 2 = Both achieved<br>1 = 1 only<br>0 = None |
| 11 | Skills | Summarizing and confirming | Summarizes and checks the accuracy of the information with the patient | 2 = Both done<br>1 = 1 only<br>0 = None |
| 12 | Skills | Closure | Checks for additional questions, makes next appointment, and expresses gratitude | 2 = All 3 done<br>1 = 1 missed<br>0 = ≥2 missed |
| 13 | Skills | Professional response | Responds appropriately as an OT when facing questions/comments | 2 = Appropriate throughout<br>1 = 1 inappropriate response<br>0 = ≥2 inappropriate |
| 14 | Evaluation | Observation reporting (appearance, expression, and posture) | Accurately reports observed physical cues during the interview | 2 = All 3 reported<br>1 = 1 missed<br>0 = ≥2 missed |
| 15 | Evaluation | Speech and thought characteristics | Reports speaking style and mental symptoms (e.g., psychotic features) | 2 = Both domains reported<br>1 = 1 only<br>0 = None |
| 16 | Evaluation | Content summary | Accurately summarizes and reports the patient's words | 2 = Both accurate<br>1 = 1 only<br>0 = None |

Scoring Criteria: The OSCE rating scale comprises three domains: attitude (0–8 points), skills (0–18 points), and evaluation (0–6 points), with a total possible score of 0–32 points. Each item is rated on a 3-point scale (0 = not demonstrated, 1 = partially demonstrated, 2 = fully demonstrated) based on observable behaviors relevant to psychiatric intake interviews.
