## Supplementary material for "Development and Reliability of an Objective Structured Clinical Examination for Intake Interviews in Mental Health Occupational Therapy Education": Demographic and professional characteristics of the expert panel involved in the development of the OSCE rating scale

**Supplementary Table 2. Demographic and professional characteristics of the expert panel involved in the development of the OSCE rating scale.**

| Characteristic | Category | n |
|---|---|---|
| Sex | Male | 5 |
| Age group(years) | 30-39 | 3 |
| | 40-49 | 2 |
| Primary role | Occupational therapy educator(mental health) | 2 |
| | Clinical occupational therapist(mental health) | 3 |
| Clinical experience(years) | 10-19 | 3 |
| | 20-29 | 2 |
| Practice setting | University | 2 |
| | Hospital or psychiatric daycare | 2 |
| | Community-based rehabilitation | 1 |

Abbreviations: OT, occupational therapist; OSCE, Objective Structured Clinical Examination.

To preserve anonymity, institution names and geographic locations are not reported.
